# Outbreaks of publications about emerging infectious diseases: the case of SARS-CoV-2 and Zika virus

**DOI:** 10.1101/2020.11.20.20235242

**Authors:** Aziz Mert Ipekci, Diana Buitrago-Garcia, Kaspar Walter Meili, Fabienne Krauer, Nirmala Prajapati, Shabnam Thapa, Lea Wildisen, Lucia Araujo Chaveron, Lukas Baumann, Sanam Shah, Tessa Whiteley, Gonzalo Solís-García, Foteini Tsotra, Ivan Zhelyazkov, Hira Imeri, Nicola Low, Michel Jacques Counotte

## Abstract

**Background:** Outbreaks of infectious diseases generate outbreaks of scientific evidence. In 2016 epidemics of Zika virus emerged, largely in Latin America and the Caribbean. In 2020, a novel severe acute respiratory syndrome coronavirus 2 (SARS-CoV-2) caused a pandemic of coronavirus disease 2019 (COVID-19). We compared patterns of scientific publications for the two infections over time.

**Methods:** We used living systematic review methods to search for and annotate publications according to study design. For Zika virus, a review team performed the tasks for publications in 2016. For SARS-CoV-2, a crowd of 25 volunteer scientists performed the tasks for publications up to May 24, 2020. We used descriptive statistics to categorise and compare study designs over time.

**Findings:** We found 2,286 publications about Zika virus in 2016 and 21,990 about SARS-CoV-2 up to 24 May 2020, of which we analysed a random sample of 5294. For both infections, there were more epidemiological than laboratory science studies. Amongst epidemiological studies for both infections, case reports, case series and cross-sectional studies emerged first, cohort and case-control studies were published later. Trials were the last to emerge. Mathematical modelling studies were more common in SARS-CoV-2 research. The number of preprints was much higher for SARS-CoV-2 than for Zika virus.

**Interpretation:** Similarities in the overall pattern of publications might be generalizable, whereas differences are compatible with differences in the characteristics of a disease. Understanding how evidence accumulates during disease outbreaks helps us understand which types of public health questions we can answer and when.

**Funding:** MJC and HI are funded by the Swiss National Science Foundation (SNF grant number 176233). NL acknowledges funding from the European Union’s Horizon 2020 research and innovation programme - project EpiPose (grant agreement number 101003688). DBG is funded by the Swiss government excellence scholarship (2019.0774) and the Swiss School of Public Health Global P3HS.

## Introduction

Scientists publish their findings to understand epidemics caused by novel pathogens. This evidence will guide decisions, actions and interventions to mitigate the effects of the disease through policy, programmes, guidelines and further research.^1^ Two viral pathogens that have caused epidemics across a large number of countries since 2016 resulted in the declaration of a Public Health Emergency of International Concern (PHEIC) by the World Health Organization (WHO) Director-General.^2^ Zika virus, a mosquito-borne virus caused epidemics of microcephaly that were first noticed in late 2015 in Brazil, although it was first discovered in 1947 and had caused small outbreaks of infection before then.^3^ Severe acute respiratory syndrome coronavirus 2 (SARS-CoV-2), was first discovered in January 2020 as the cause of a new zoonotic disease, coronavirus disease 2019 (COVID-19), spread primarily through the respiratory route.^4^ There are marked differences in the natural history of the two diseases, where microcephaly caused by Zika virus infection only emerges months after infection, COVID-19 occurs acutely. Intensive research efforts for both infections were catalysed by the needs of national, regional and global health agencies to answer key questions on transmission, prevention, and interventions at the individual and community level.^5^ During the SARS-CoV-2 pandemic, the accumulation of peer-reviewed and preprint publications has been vast; from April, 2020 onwards, an average of 2,000 scientific publications appeared per week.^6^ A similar, albeit smaller surge in publications occurred in 2016 during the Zika virus epidemic. The sudden large increases in publications about these conditions over a short time can also be described as outbreaks.

The emergence of a new disease provides an opportunity to examine how research evidence emerges and develops, according to the research question and the feasibility of the study methods. Hierarchies of evidence are often used to rank the value of epidemiological study designs, prioritising experimental methods,^7^ but these do not take account of purposes, other than the effects of interventions. Anecdotal observations allow for the discovery and description of phenomena, studies with comparison groups are more appropriate to test hypotheses, and randomised trials test the causal effects of interventions.^8^ Early on in the Zika epidemic, questions about causality were important because the link between clusters of babies born with microcephaly and Zika virus infection was not obvious; congenital abnormalities caused by a mosquito-borne virus had never been reported. In an analysis of 346 publications about Zika virus, we described the temporal sequence of publication of types of study to investigate causality.^9^ Others have assessed the accumulation of study designs over time during the SARS-CoV-2 outbreak and concluded that early in the outbreak, simple observational studies and narrative reviews were most abundant.^10^ Here, we proposed a hypothetical sequence: first, anecdotal observations are reported in case reports or case series. Analytical observational studies follow. In parallel, basic research studies investigate the biology and pathogenesis of the disease. Mathematical modelling can provide evidence where direct observations are not available.^11^ After a delay, controlled trials examining interventions are published.

The emergence of SARS-CoV-2 allows a comparison with Zika virus between the timing and types of evidence published at the start of an outbreak of a new disease. The objectives of this study were to analyse the patterns of evolution of the evidence over time during the 2016 Zika virus epidemic and the 2020 SARS-CoV-2 pandemic. We compare the sequence of evidence accumulation with the previously hypothesised pattern.^9^

## Methods

### Data collection

#### Searches and sources

We used databases that were created for the Zika Open Access Project (ZOAP) and COVID-19 Open Access Project (COAP).^12^ For each pathogen, we ran daily automated searches to index and deduplicate records of articles about Zika virus (from January 1, 2016) and SARS-CoV-2 research (from January 1, 2020) in EMBASE via OVID, MEDLINE via PubMed, and the preprint server bioRxiv (for SARS-CoV-2 we also searched medRxiv). We specify the search terms in the appendix (pp 2).

#### Annotation of records with study design

We screened the title and abstract, or full text when the first was insufficient, and annotated each record with its study design. For weeks where the volume SARS-CoV-2 of research was over 400 publications, starting mid-March, we drew a random sample of 400 publications. The annotation of the Zika virus dataset was performed (from January 1, 2016 to December 31, 2016) for previous systematic reviews.^13, 14^

#### Study design classification

We first classified publications into the broad groups “epidemiology”, “basic research”, “non-original research” (editorials, viewpoints, and commentaries) and “other”. We subdivided epidemiological and basic research further, based on their study design. We provide details on the classification of the study designs in the appendix (pp 3).

#### Crowd

To distribute the annotation workload, we recruited a ‘crowd’ of volunteer scientists.^15^ We included researchers with a background in medicine or public health, who qualified by passing a pilot test using an online tool that simulates classification tasks. A demonstration and the source code of the tool are provided online.^16^

The crowd members used another online tool for screening, annotation, and verification of each record. A first crowd member screened and annotated a record, and a second crowd member verified the annotated data. Disagreements were resolved by a third member of the team. One person (MJC) distributed tasks centrally and a ‘crowd supervisor’ (AMI) monitored progress. Crowd members took part in the interpretation of the results.

#### Reported number of cases

To compare the number of publications against the number of reported cases, we used open-source data on Zika virus and SARS-CoV-2 from, see Data sharing.

#### Date that a publication becomes available

We defined the date at which a publication became available as the date it was indexed in the MEDLINE or EMBASE database, or when it appeared on the preprint server.

### Data analysis

First, we described the evolution of reported cases and publications over time. Second, we described the proportions of study designs, by week, for SARS-CoV-2 and by month for Zika virus, due to the differences in research volume. We omitted the first two weeks of 2020 for SARS-CoV-2 because there were only four publications, making the proportions unstable. To take into account the random sampling of the SARS-CoV-2 research, we provided the Wilson score 95% confidence intervals (CI) for the proportions. Third, we quantified the timing and speed of the accumulation of publications of different study designs: We plotted the time elapsed between the first and twentieth occurrence of publications of each study design. Last, we described the proportion of evidence that was published on preprint servers during the two epidemics, and by study design.

## Results

Between week one and week 21 (up to May 24) 2020, we indexed 21,990 publications, and a crowd of 25 contributors annotated a sample of 5,294 publications on SARS-CoV-2. For the Zika virus research, we annotated all 2,286 identified publications for 2016. Both the volume of the weekly reported cases and number of publications were 30-50-fold higher for SARS-CoV-2 than for Zika virus (figure 1).

**Figure 1.**
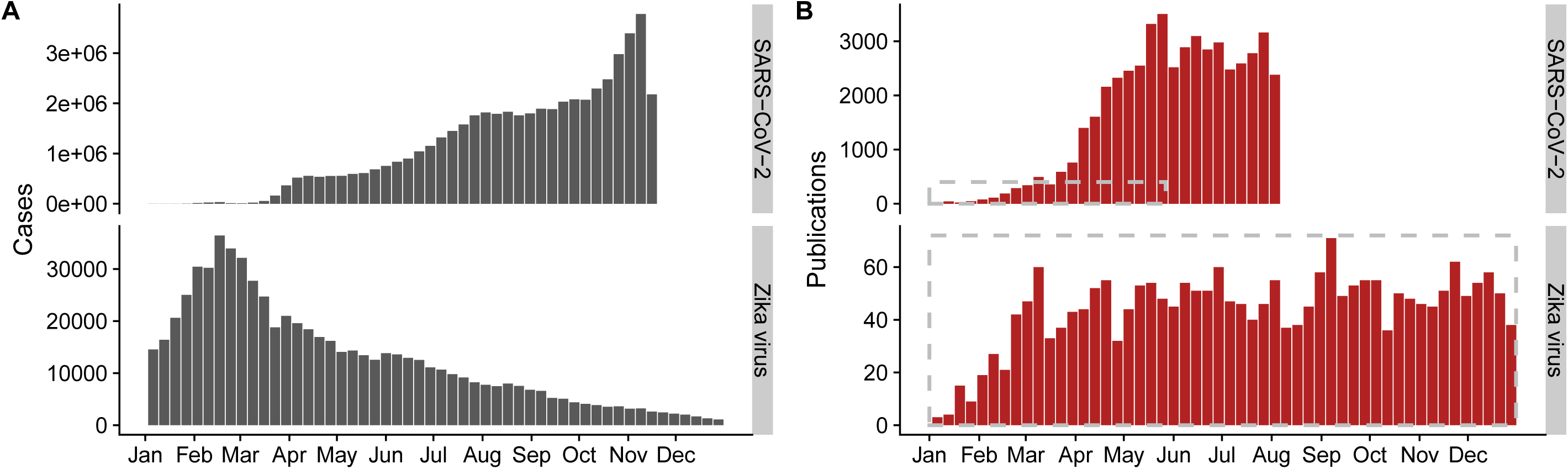
The global number of reported cases (A), and the number publications (B) by week for SARS-CoV-2 infections in 2020 and Zika virus infections (ZIKV) in 2016. In panel B, the dashed grey boxes contain the period and number of publications for which the study design was annotated. The vertical scales differ for each infection. SARS-CoV-2: Severe acute respiratory syndrome coronavirus 2.

### The proportion of different study designs

In both epidemics, a substantial and reasonably stable proportion of the publications were non-original research. The overall proportion of non-original publications was higher for Zika virus (55%, (appendix pp 4)) than for SARS-CoV-2 (34% [95% CI: 33-35], (appendix pp 5)). For publications of original research, the proportion of basic research publications increased over time for Zika virus, but decreased for SARS-Cov-2 research (figure 2A).

**Figure 2.**
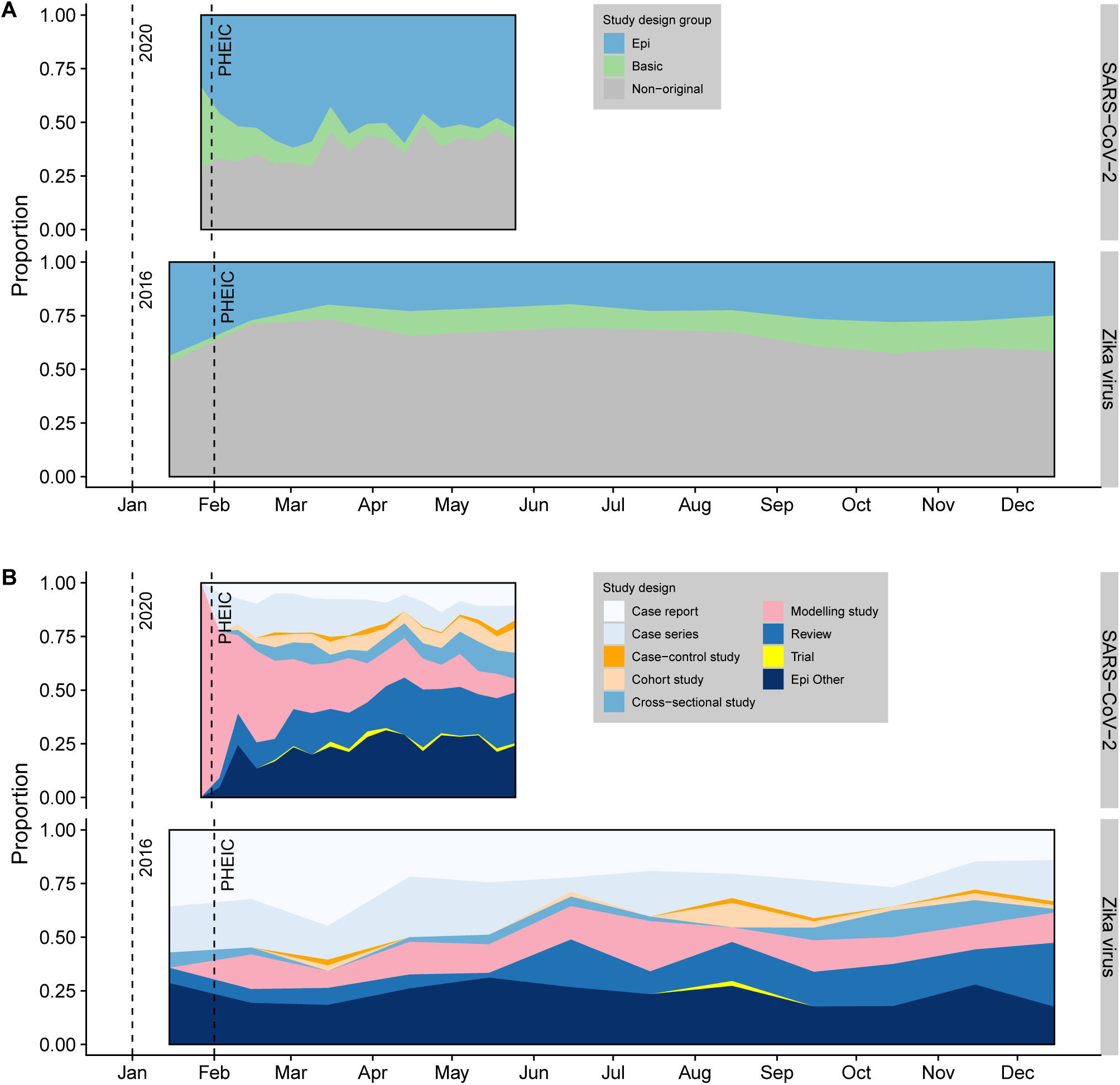
Proportions of different study designs of published research on SARS-CoV-2 (SARS-CoV-2) and Zika virus (Zika virus) over time. Epidemiological, basic, and “non-original” research (A); epidemiological research by study design (B). For display purposes SARS-CoV-2 data is shown by week and Zika virus data by month. SARS-CoV-2: Severe acute respiratory syndrome coronavirus 2; PHEIC: Public Health Emergency of International Concern.

Within the epidemiological study designs, mathematical modelling studies had a larger role at the beginning of the SARS-CoV-2 pandemic (10·1%, [95% CI: 9·3-11·0]) and compared to the Zika virus outbreak (3·2%). Many of these were published as preprint publications. When we excluded preprint publications, the evolution of evidence over time became more similar between the two epidemics (appendix pp 6). Case reports and case series accounted for approximately 10% of the total body of evidence; 10·7% [95% CI: 9·9-11·6] for SARS-CoV-2 and 9·7% for Zika virus research. Analytical epidemiological study designs became more prevalent later in the SARS-Cov-2 and Zika virus epidemics. Case-control and cohort studies accounted for 4·0% [95% CI: 3·5-4·6] for SARS-CoV-2 and 0·8% for Zika virus. Trials also emerged later but in smaller numbers (27/5,294 for SARS-CoV-2, and 1/2,286 for Zika virus) (Fig 2B).

### Accumulation of epidemiological and basic research

Despite the difference in volume, the accumulation of study designs over time for SARS-CoV-2 and Zika virus research show some similarities (figure 3). Case reports, case series and cross-sectional studies were the first epidemiological study designs to be reported, together with non-original articles and reviews. Case-control and cohort studies followed later; this delay was more prominent in the Zika virus research. Phylogenetic studies and mathematical modelling studies had a more prominent role early on during the SARS-CoV-2 pandemic than in the Zika virus epidemic. In vivo and in vitro laboratory studies followed between case reports and controlled observational studies. Trials were the last type of study to be published.

**Figure 3.**
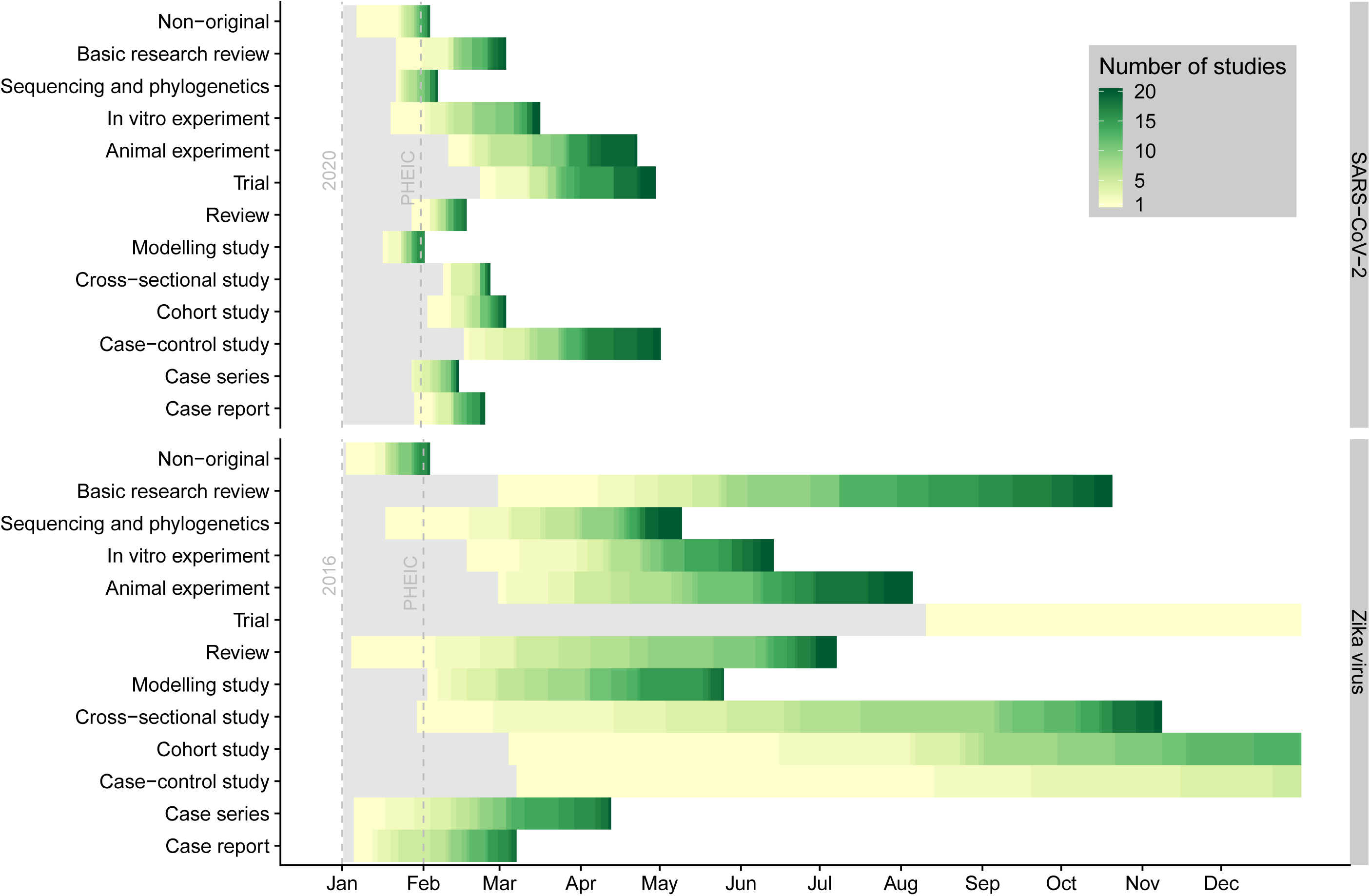
Time to the first 20 publications in a study design, for SARS-CoV-2 infections (SARS-CoV-2) and Zika virus infections (Zika virus). SARS-CoV-2: Severe acute respiratory syndrome coronavirus 2; PHEIC: Public Health Emergency of International Concern.

### The role of preprint publications

The role of preprint publications was more prominent at the start of the SARS-CoV-2 pandemic than the Zika virus epidemic. In January and February 2020, the majority of publications on SARS-CoV-2 were manuscripts on preprint servers (figure 4A). Basic research reviews were seldom published on preprint servers, whereas 77% of the mathematical modelling studies were initially made available on preprint servers (figure 4B). The proportion of modelling and sequencing studies that were published as preprints was high throughout the first 21 weeks of 2020, whereas other designs reduced over time (appendix pp 8). The proportion of preprints decreased over time.

**Figure 4.**
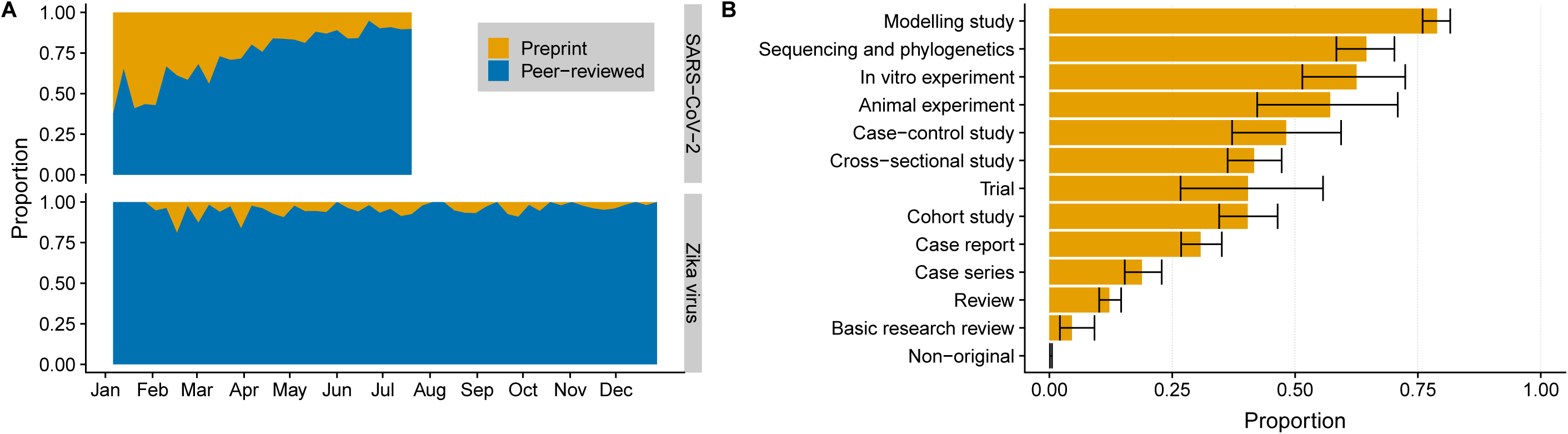
The proportion of preprint publications and peer-reviewed publications for SARS-CoV-2 (SARS-CoV-2) and Zika virus (Zika virus) research over time (A) and by study design for SARS-CoV-2 (B). SARS-CoV-2: Severe acute respiratory syndrome coronavirus 2

## Discussion

The overall distribution of publications at the start of the SARS-CoV-2 and Zika virus epidemics was similar. Epidemiological research was more commonly published than laboratory research and non-original contributions accounted for a substantial fraction of all publications for both infections. For both infections, case reports and case series, mathematical modelling and phylogenetic studies were prominent at the start of the epidemic, whereas analytical study designs, such as cohort and case-control studies, appeared later. Trials emerged later and accounted for a small proportion of all studies. The volume and speed of evolution were much higher for SARS-CoV-2 than for Zika virus. Modelling studies were more prominent and basic research studies were less common for SARS-CoV-2 than for Zika virus. More studies were published as preprints for SARS-CoV-2, but this proportion declined over time.

Strengths and limitations. Strengths of this study include the comparable and reproducible search strategies for two emerging infectious diseases and categorisation of study design by a volunteer crowd of epidemiologist reviewers. A limitation is that the design of an epidemiological study is not always clear, and different scientists might classify the same study differently. We tried to tackle this limitation by screening and training of the volunteer scientists, verification of decisions and having a third person resolving disagreements.^12^ There are other limitations. First, we only recorded the study design of publications and did not assess the content or its methodological quality. To trace the evolution of evidence for specific research questions, in-depth studies are needed. Second, for SARS-CoV-2, the volume of publications meant that we only annotated a sample of records. The total in the first five months of the pandemic was, however, higher than for one year of publications about Zika virus and the proportions of different study designs for Zika virus stabilised quickly. Third, the searches do not include all sources of peer-reviewed evidence or preprint sources. Incompleteness of the evidence base should not affect our conclusions as long as other sources account for a stable proportion of publications.

We followed two dimensions of the publication of evidence about two newly emerging infectious diseases; the overall distribution of publication types and changes over time. Similarities in the overall distribution of epidemiological, basic science and non-original publications for SARS-CoV-2 and Zika virus could reflect patterns of the overall trajectory of research about emerging infectious diseases. In the initial phase of an outbreak with a novel pathogen, case reports and case series predominate. These types of study describe and refine the clinical characteristics of the disease.^17^ Observations from these studies are commonly used to define research questions and formulate hypotheses about various aspects of transmission and disease. More formal, hypothesis-driven and interventional research follows later.^8^

The differences between study designs in the two epidemics are compatible with differences in characteristics of the diseases. The higher proportion of basic research in Zika virus research may have several explanations. First, the occurrence of congenital abnormalities following a vector-borne infection was poorly understood; in vivo and studies were essential to investigate in utero transmission and mechanisms for neurotoxicity and neuropathology.^18^ Second, the establishment of mouse models was more successful in Zika virus research than for SARS-CoV-2 research,^19^ although efforts are ongoing.^20^ Third, the later occurrence of case-control studies and cohorts studies in Zika virus, might be caused by the delay to congenital outcomes, compared to the shorter delay in outcomes caused by SARS-CoV-2. Fourth, the prominent role of mathematical modelling studies during the beginning of the SARS-CoV-2 pandemic, probably reflects early recognition of the pandemic potential and the need for forecasts of the global spread. Mechanistic models describing the transmission SARS-CoV-2 are also less complex than for arboviruses like Zika virus, allowing many to explore transmission dynamics.^11^ The higher volume of observational research about SARS-CoV-2 research could reflect both the 50-fold higher numbers of cases than for Zika virus and the severity of the pandemic, whereas Zika virus was largely limited to the Americas and cases of infection were already declining as the research volume started to increase. The increasing role of preprints during the SARS-CoV-2 coincided with developments in open access publishing and the need for speedy access to outbreak research.^21^

Other researchers have studied the evolution of evidence during disease outbreaks as well. During the SARS outbreak in 2003, Xing et al. (2010) described epidemiological studies about Toronto and Hong Kong, whereas we included epidemiological and non-epidemiological articles all over the globe.^22^ Xing and colleagues primarily studied the publication time delay during the outbreak and concluded that only a minority (7%) of the publications was published during the time outbreak, while we investigated the proportion of the preprints over peer-reviewed publications.^22^ For the SARS-CoV-2 pandemic, Liu et al. (2020) performed a bibliometric analysis of the SARS-CoV-2 literature up to March 24, 2020,^23^ classifying research by theme, rather than by study design. They observed that clinical features of the COVID-19 were studied heavily, whereas other research areas such as mental health, the use of novel technologies and artificial intelligence, and pathophysiology remained underexplored. In contrary to the manual annotation of our project, Tran et al. (2020) performed automatic Latent Dirichlet allocation topic modelling of publications on SARS-CoV-2, published up to April 23, 2020,^24^ with findings similar to those of Liu. et al. While we validated classification of study design manually, Tran et al. did not describe a validation of their automated modelling method. Jones et al. (2020) showed a similar pattern of study design occurrence during the early SARS-CoV-2 pandemic, where case reports and narrative reviews were found to be most published.^10^ However, they merely present absolute numbers and a comparison with other outbreaks is absent.^10^ Haghani and Bliemer (2020) compared SARS, MERS and SARS-CoV-2 literature and showed that around 50% of studies were non-original, which is in line with our results.^2^5 Unlike our categorization method, Haghani and Bliemer used the categorization by the citation database ‘SCOPUS’ and conclude that the studies linked to public health response are first to emerge.^25^

Our work has several implications for policy and research. The change over time in the types of studies has particular implications for synthesis of evidence as more research is published. The earliest publications about a new pathogen come from studies that can be done quickly and non-original commentaries. The earliest studies published might not be the most appropriate to answer specific questions, for example, about causality,^26^ or to quantify disease characteristics. Triangulation of different sources of evidence using frameworks, such as those based on the Bradford Hill criteria,^13^ and careful interpretation through explicit acknowledgment of limitations can help. Living systematic reviews are particularly useful because changes to inclusion criteria can be planned and protocols can be amended in advance of an update. For example, quantifying the proportion of asymptomatic SARS-CoV-2 infections in March 2020 relied largely on descriptions from contact investigations in single families.^27^ By June 2020, there were also studies at lower risk of selection and measurement biases, such as population screening. ^28^

The vast quantity of evidence about emerging infections poses challenges for the efficient handling and evaluation of information. The speed with which the evidence about SARS-CoV-2 has accumulated is unprecedented. We recruited a large team of experienced scientists, but we were still not able to categorise all publications by the time this manuscript is written. Machine learning methods, such as natural language processing, to classify text is a promising approach for the triage of publication types.^15^ We also see potential in a scaled-up version of collaborative crowd-sourcing among experts in the field, to increase efficiency and avoid research waste.^29^ The technical tools to manage such efforts are available, but guidelines on how to best conduct the live synthesis of evidence should be developed and evaluated further.^30^ The findings of this study show how description of the types and timing of publications during outbreaks of emerging and re-emerging diseases can help us understand which types of public health questions we can answer and when. Further analyses of the generation and accumulations of research evidence during disease outbreaks could help to improve the public health response.

## Data Availability

All data is available online.

https://doi.org/10.7910/DVN/IPHUJN

## Contributions

**Conceptualization:**MJC, NL; **Methodology:** MJC, NL, AMI; **Data collection:** AMI, DBG, KWM, FK, NP, ST, LW, LAC, LB, SS, TW, GSG, FT, IZ, HI, NL, MJC; **Analyses:** MJC, AMI; AMI, DBG, KWM, FK, NP, ST, LW, LAC, LB, SS, TW, GSG, FT, IZ, HI, NL, MJC; **Interpretation of analyses: preparation:** MJC, AMI; **Writing – review and editing:** AMI, DBG, KWM, FK, NP, ST, LW, LAC, LB, SS, TW, GSG, FT, IZ, HI, NL, MJC; **Supervision:** MJC, NL;

## Data sharing

All data is available online: https://doi.org/10.7910/DVN/IPHUJN

## References

1. Brownson RC, Fielding JE, Maylahn CM. Evidence-based public health: a fundamental concept for public health practice. 2009; 30: 175–201.

2. World Health Organization. WHO statement on the first meeting of the International Health Regulations (2005) (IHR 2005) Emergency Committee on Zika virus and observed increase in neurological disorders and neonatal malformations. 2016. https://www.who.int/news-room/detail/01-02-2016-who-statement-on-the-first-meeting-of-the-international-health-regulations-(2005)-(ihr-2005)-emergency-committee-on-zika-virus-and-observed-increase-in-neurological-disorders-and-neonatal-malformations (accessed 10/10/2020).

3. World Health Organization. COVID-19 Public Health Emergency of International Concern (PHEIC) Global research and innovation forum. 2020. https://www.who.int/publications/m/item/covid-19-public-health-emergency-of-international-concern-(pheic)-global-research-and-innovation-forum (accessed 10/10/2020).

4. Wiersinga WJ, Rhodes A, Cheng AC, Peacock SJ, Prescott HC. Pathophysiology, Transmission, Diagnosis, and Treatment of Coronavirus Disease 2019 (COVID-19): A Review. JAMA 2020; 324(8): 782–93.

5. Zhang L, Zhao W, Sun B, Huang Y, Glanzel W. How scientific research reacts to international public health emergencies: a global analysis of response patterns. Scientometrics 2020; 124(1): 1–27.

6. Counotte MJ, Imeri H, Ipekci M, Low N. Living Evidence on COVID-19. 2020. https://ispmbern.github.io/covid-19/living-review/ (accessed 31/07/2020).

7. Borgerson K. Valuing evidence: bias and the evidence hierarchy of evidence-based medicine. Perspectives in biology and medicine 2009; 52(2): 218–33.

8. Vandenbroucke JP. Observational research, randomised trials, and two views of medical science. PLoS medicine 2008; 5(3): e67.

9. Counotte MJ, Meili KW, Low N. Emergence of evidence during disease outbreaks: lessons learnt from the Zika virus outbreak. medRxiv 2020: 2020.03.16.20036806.

10. Jones RC, Ho JC, Kearney H, et al. Evaluating Trends in COVID-19 Research Activity in Early 2020: The Creation and Utilization of a Novel Open-Access Database. Cureus 2020; 12(8): e9943.

11. Metcalf CJE, Morris DH, Park SW. Mathematical models to guide pandemic response. Science 2020; 369(6502): 368–9.

12. COVID-19 Open Acces Project. Screening and annotation of citations. 2020. https://ispmbern.github.io/covid-19/living-review/screening.html (accessed 07/08/2020).

13. Krauer F, Riesen M, Reveiz L, et al. Zika Virus Infection as a Cause of Congenital Brain Abnormalities and Guillain-Barre Syndrome: Systematic Review. PLoS medicine 2017; 14(1): e1002203.

14. Counotte MJ, Egli-Gany D, Riesen M, et al. Zika virus infection as a cause of congenital brain abnormalities and Guillain-Barre syndrome: From systematic review to living systematic review. F1000Research 2018; 7: 196.

15. Thomas J, Noel-Storr A, Marshall I, et al. Living systematic reviews: 2. Combining human and machine effort. Journal of clinical epidemiology 2017; 91: 31–7.

16. Counotte MJ. Rapid screening with a R Shiny App. 2020. https://github.com/ZikaProject/ShinyScreeningExample (accessed 03-08-2020.

17. Vandenbroucke JP. In defense of case reports and case series. Annals of internal medicine 2001; 134(4): 330–4.

18. Morrison TE, Diamond MS. Animal Models of Zika Virus Infection, Pathogenesis, and Immunity. Journal of virology 2017; 91(8).

19. Lakdawala SS, Menachery VD. The search for a COVID-19 animal model. Science 2020; 368(6494): 942–3.

20. Sun SH, Chen Q, Gu HJ, et al. A Mouse Model of SARS-CoV-2 Infection and Pathogenesis. Cell host & microbe 2020; 28(1): 124–33 e4.

21. Kupferschmidt K. ‘A completely new culture of doing research.’ Coronavirus outbreak changes how scientists communicate. Science 2020.

22. Xing W, Hejblum G, Leung GM, Valleron AJ. Anatomy of the epidemiological literature on the 2003 SARS outbreaks in Hong Kong and Toronto: a time-stratified review. PLoS medicine 2010; 7(5): e1000272.

23. Liu N, Chee ML, Niu C, et al. Coronavirus disease 2019 (COVID-19): an evidence map of medical literature. BMC medical research methodology 2020; 20(1): 177.

24. Tran BX, Ha GH, Nguyen LH, et al. Studies of Novel Coronavirus Disease 19 (COVID-19) Pandemic: A Global Analysis of Literature. International journal of environmental research and public health 2020; 17(11).

25. Haghani M, Bliemer MCJ. Covid-19 pandemic and the unprecedented mobilisation of scholarly efforts prompted by a health crisis: Scientometric comparisons across SARS, MERS and 2019-nCoV literature. Scientometrics 2020: 1–32.

26. Higgins JP, Ramsay C, Reeves BC, et al. Issues relating to study design and risk of bias when including non-randomized studies in systematic reviews on the effects of interventions. Research synthesis methods 2013; 4(1): 12–25.

27. Buitrago-Garcia DC, Egli-Gany D, Counotte MJ, et al. The role of asymptomatic SARS-CoV-2 infections: rapid living systematic review and meta-analysis [Version 1]. medRxiv 2020: 2020.04.25.20079103.

28. Buitrago-Garcia D, Egli-Gany D, Counotte MJ, et al. Occurrence and transmission potential of asymptomatic and presymptomatic SARS-CoV-2 infections: A living systematic review and meta-analysis. PLoS medicine 2020; 17(9): e1003346.

29. Macleod MR, Michie S, Roberts I, et al. Biomedical research: increasing value, reducing waste. Lancet 2014; 383(9912): 101–4.

30. Khamis AM, Kahale LA, Pardo-Hernandez H, Schunemann HJ, Akl EA. Methods of conduct and reporting of living systematic reviews: a protocol for a living methodological survey. F1000Research 2019; 8: 221.

