## Supplementary Material for "Outbreaks of publications about emerging infectious diseases: the case of SARS-CoV-2 and Zika virus"

**Supplementary Text 1. Search strategy**
**COVID:**

EMBASE:

(SARS coronavirus/ OR middle east respiratory syndrome/ OR severe acute respiratory syndrome/ OR (coronavirus* OR corona virus* OR HCoV* or ncov* OR covid or covid19 OR sars-cov* or sarscov* or Sars-coronavirus* OR Severe Acute Respiratory Syndrome Coronavirus*).mp.) and 20191201:20301231.(dc).

MEDLINE:

("coronavirus"[MH] OR "coronavirus infections"[MH] OR "coronavirus"[TW] OR "corona virus"[TW] OR "HCoV"[TW] OR "nCov"[TW] OR "covid"[TW] OR "covid19"[TW] OR "Severe Acute Respiratory Syndrome Coronavirus 2"[TW] OR "SARS-CoV2"[TW] OR "SARS-CoV 2"[TW] OR "SARS Coronavirus 2"[TW] OR "MERS-CoV"[TW]) AND (2019/1/1:3000[PDAT])

From 01.04.2020, we retrieve the currate BioRxiv/MedRxiv dataset [https://connect.medrxiv.org/relate/content/181 ]

https://ispmbern.github.io/covid-19/living-review/collectingdata.html

**Zika:** All sources: Zika OR ZIKV OR “Zika virus”

**Supplementary Table S1. Classification of study designs, and the group and original status to which we assigned studies.**

| Label | Code | Group |
|---|---|---|
| Case report | 1 | Epi |
| Case series | 2 | Epi |
| Case-control study | 3 | Epi |
| Cohort study | 4 | Epi |
| Cross-sectional study | 5 | Epi |
| Diagnostic study | 6 | Epi |
| Ecological study | 7 | Epi |
| Guidelines | 8 | Epi |
| Modelling study | 9 | Epi |
| Other | 10 | Epi |
| Outbreak or surveillance report | 11 | Epi |
| Qualitative study | 12 | Epi |
| Review | 13 | Epi |
| Trial | 14 | Epi |
| Animal experiment | 21 | Basic |
| In vitro experiment | 22 | Basic |
| Biochemical/protein structure studies | 27 | Basic |
| Sequencing and phylogenetics | 23 | Basic |
| Within-host modelling | 24 | Basic |
| Basic research review | 25 | Basic |
| Other | 26 | Other |
| Comment/editorial/non-original | 30 | Non-original |

Epi: epidemiological study designs; Basic: basic research study design; Other: classified as neither Epi nor Basic research.

**Supplementary Table S2. Number and proportion of publications on Zika virus by category and publication type.**

| | **Preprint (N=98)** | **Peer-reviewed (N=2188)** | **Overall (N=2286)** |
|---|---|---|---|
| **Study design group** | | | |
| Epi | 31 (31·6%) | 521 (23·8%) | 552 (24·1%) |
| Basic | 37 (37·8%) | 215 (9·8%) | 252 (11·0%) |
| Other | 30 (30·6%) | 1452 (66·4%) | 1482 (64·8%) |
| **Source** | | | |
| MedRxiv | 0 (0%) | 0 (0%) | 0 (0%) |
| BioRxiv | 98 (100%) | 0 (0%) | 98 (4·3%) |
| EMBASE | 0 (0%) | 356 (16·3%) | 356 (15·6%) |
| Pubmed | 0 (0%) | 1832 (83·7%) | 1832 (80·1%) |
| **Study design** | | | |
| Case report | 1 (1·0%) | 128 (5·9%) | 129 (5·6%) |
| Case series | 2 (2·0%) | 92 (4·2%) | 94 (4·1%) |
| Case-control study | 0 (0%) | 5 (0·2%) | 5 (0·2%) |
| Cohort study | 0 (0%) | 13 (0·6%) | 13 (0·6%) |
| Cross-sectional study | 0 (0%) | 27 (1·2%) | 27 (1·2%) |
| Diagnostic study | 0 (0%) | 30 (1·4%) | 30 (1·3%) |
| Ecological study | 3 (3·1%) | 9 (0·4%) | 12 (0·5%) |
| Guidelines | 0 (0%) | 22 (1·0%) | 22 (1·0%) |
| Modelling study | 22 (22·4%) | 52 (2·4%) | 74 (3·2%) |
| Other | 6 (6·1%) | 37 (1·7%) | 43 (1·9%) |
| Outbreak or surveillance report | 0 (0%) | 33 (1·5%) | 33 (1·4%) |
| Qualitative study | 0 (0%) | 23 (1·1%) | 23 (1·0%) |
| Review | 0 (0%) | 82 (3·7%) | 82 (3·6%) |
| Trial | 0 (0%) | 1 (0·0%) | 1 (0·0%) |
| Animal experiment | 8 (8·2%) | 45 (2·1%) | 53 (2·3%) |
| In vitro experiment | 8 (8·2%) | 79 (3·6%) | 87 (3·8%) |
| Biochemical/protein structure studies | 10 (10·2%) | 62 (2·8%) | 72 (3·1%) |
| Sequencing and phylogenetics | 21 (21·4%) | 57 (2·6%) | 78 (3·4%) |
| Within-host modelling | 0 (0%) | 0 (0%) | 0 (0%) |
| Basic research review | 0 (0%) | 34 (1·6%) | 34 (1·5%) |
| Non-original | 8 (8·2%) | 1248 (57·0%) | 1256 (54·9%) |
| Mosquito | 9 (9·2%) | 109 (5·0%) | 118 (5·2%) |

**Supplementary Table S3. Number and proportion of publications on SARS-CoV-2 by category and publication type.**

| | Preprint (N=1244) | Peer-reviewed (N=4050) | Overall (N=5294) |
|---|---|---|---|
| **Study design group** | | | |
| Epi | 909 (73·1%) | 1874 (46·3%) | 2783 (52·6%) |
| Basic | 194 (15·6%) | 218 (5·4%) | 412 (7·8%) |
| Other | 141 (11·3%) | 1958 (48·3%) | 2099 (39·6%) |
| **Source** | | | |
| MedRxiv | 950 (76·4%) | 0 (0%) | 950 (17·9%) |
| BioRxiv | 294 (23·6%) | 0 (0%) | 294 (5·6%) |
| EMBASE | 0 (0%) | 676 (16·7%) | 676 (12·8%) |
| Pubmed | 0 (0%) | 3374 (83·3%) | 3374 (63·7%) |
| **Study design** | | | |
| Case report | 5 (0·4%) | 231 (5·7%) | 236 (4·5%) |
| Case series | 67 (5·4%) | 264 (6·5%) | 331 (6·3%) |
| Case-control study | 13 (1·0%) | 24 (0·6%) | 37 (0·7%) |
| Cohort study | 70 (5·6%) | 105 (2·6%) | 175 (3·3%) |
| Cross-sectional study | 94 (7·6%) | 125 (3·1%) | 219 (4·1%) |
| Diagnostic study | 73 (5·9%) | 61 (1·5%) | 134 (2·5%) |
| Ecological study | 36 (2·9%) | 24 (0·6%) | 60 (1·1%) |
| Guidelines | 3 (0·2%) | 264 (6·5%) | 267 (5·0%) |
| Modelling study | 413 (33·2%) | 122 (3·0%) | 535 (10·1%) |
| Other | 45 (3·6%) | 166 (4·1%) | 211 (4·0%) |
| Outbreak or surveillance report | 23 (1·8%) | 78 (1·9%) | 101 (1·9%) |
| Qualitative study | 4 (0·3%) | 25 (0·6%) | 29 (0·5%) |
| Review | 66 (5·3%) | 475 (11·7%) | 541 (10·2%) |
| Trial | 17 (1·4%) | 10 (0·2%) | 27 (0·5%) |
| Animal experiment | 17 (1·4%) | 14 (0·3%) | 31 (0·6%) |
| In vitro experiment | 40 (3·2%) | 21 (0·5%) | 61 (1·2%) |
| Biochemical/protein structure studies | 116 (9·3%) | 66 (1·6%) | 182 (3·4%) |
| Sequencing and phylogenetics | 118 (9·5%) | 65 (1·6%) | 183 (3·5%) |
| Within-host modelling | 14 (1·1%) | 2 (0·0%) | 16 (0·3%) |
| Basic research review | 5 (0·4%) | 116 (2·9%) | 121 (2·3%) |
| Non-original | 5 (0·4%) | 1792 (44·2%) | 1797 (33·9%) |

**Supplementary Figure S1. Proportion of different study designs of published research on SARS-CoV-2 (SARS-CoV-2) and Zika virus (ZIKV) over time, without preprint publications.**

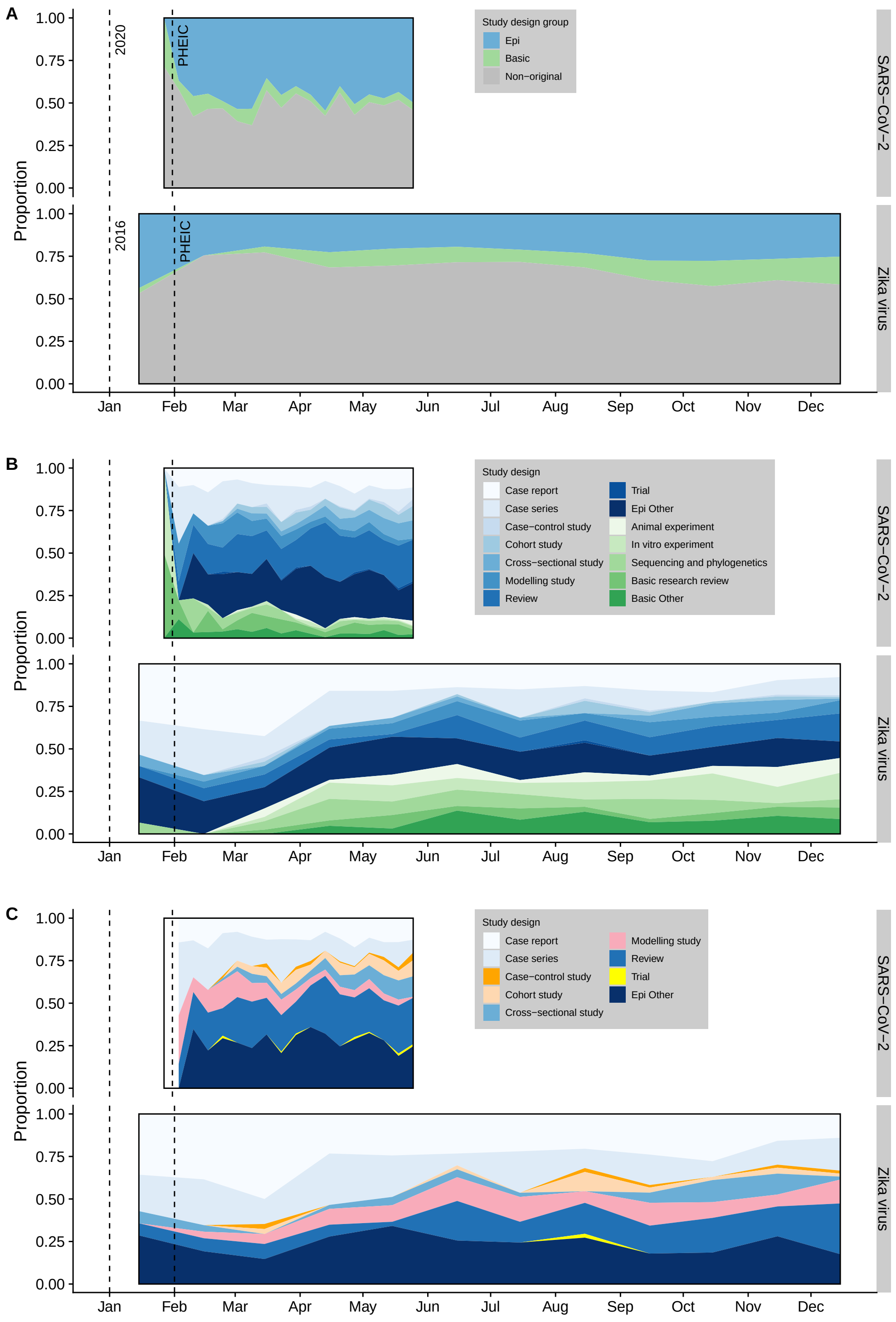

Epidemiological (Epi), basic research and non-original research (A); Epidemiological and basic research by study design (B); epidemiological research by study design (C). PHEIC: Public Health Emergency of International Concern.

**Supplementary Figure S2. Number of different study designs of published research on SARS-CoV-2 (SARS-CoV-2) and Zika virus (ZIKV) over time.**

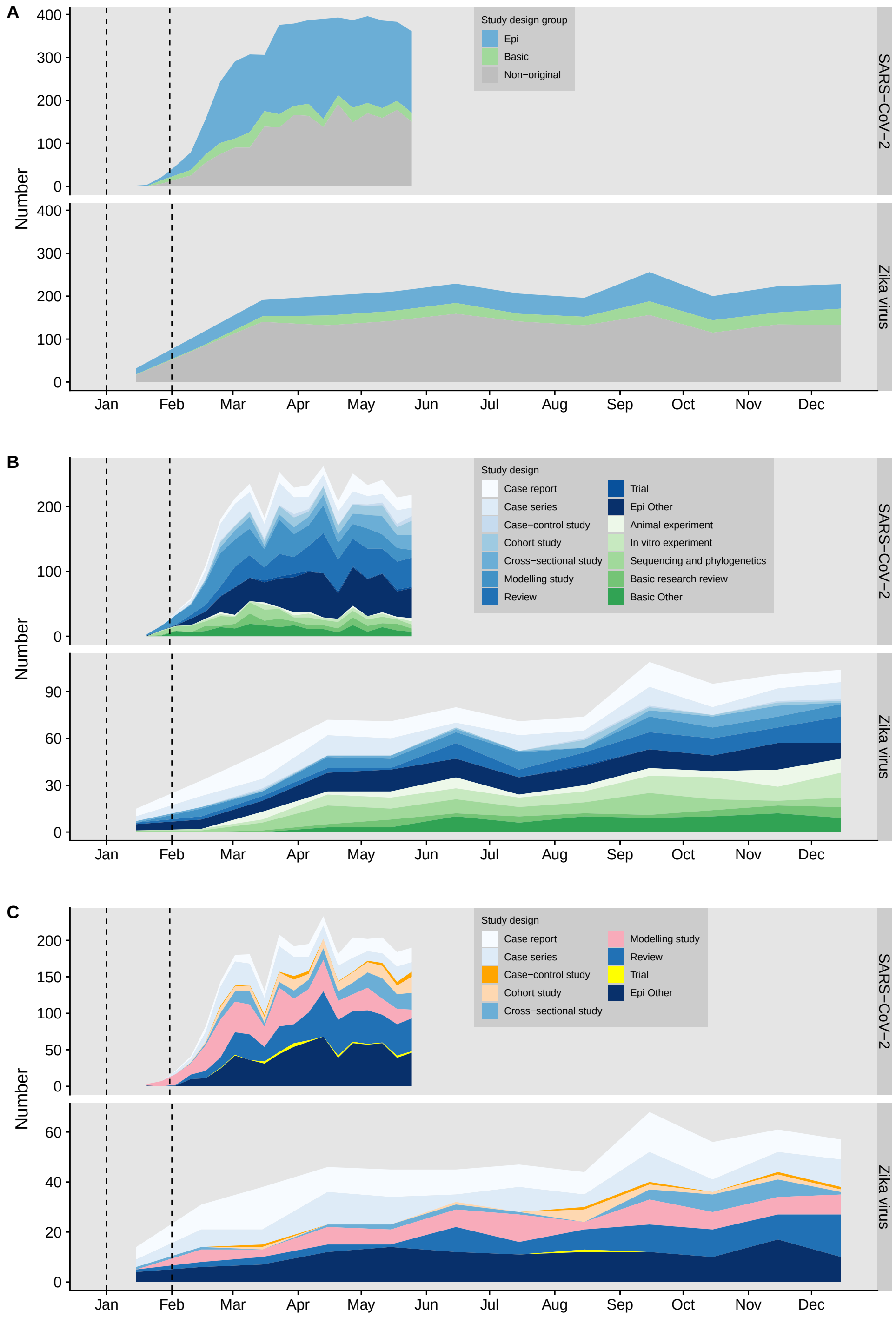

Epidemiological (Epi), basic research and non-original research (A); Epidemiological and basic research by study design (B); epidemiological research by study design (C). PHEIC: Public Health Emergency of International Concern.

**Supplementary Figure S3. The proportion of preprint publications compared to and peer-reviewed publications for SARS-CoV-2 by study design and week.**

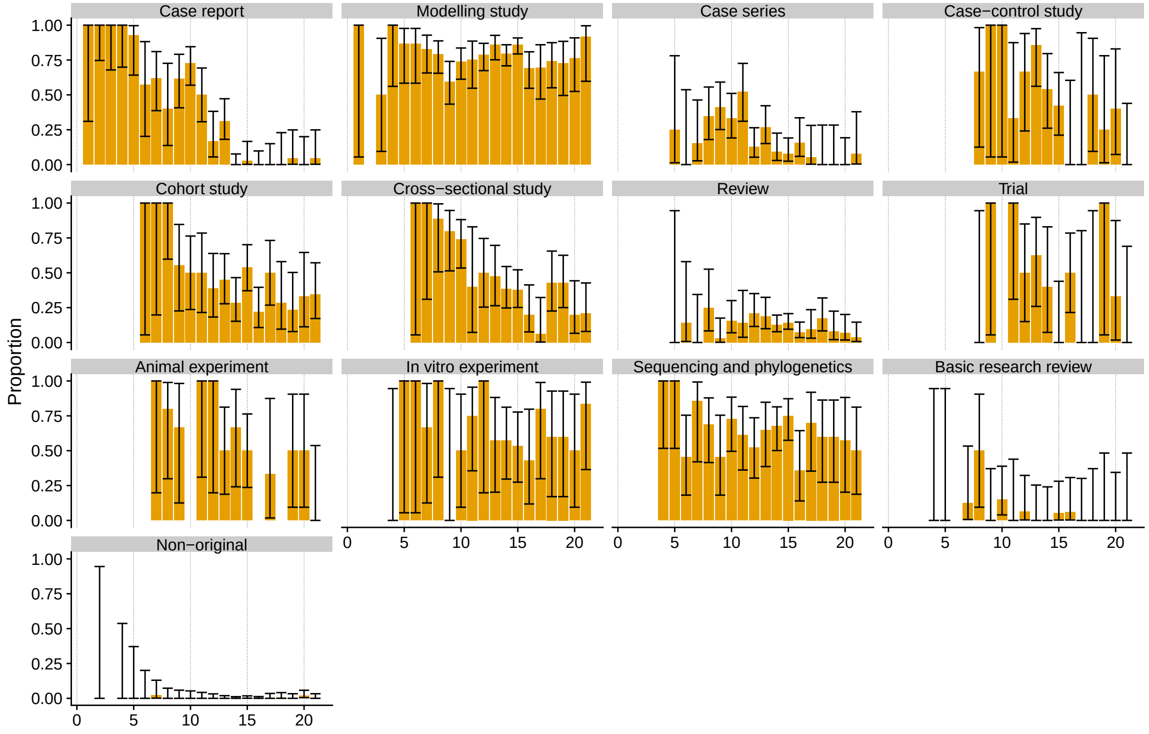
